# A prospective study of the METS-IR index to predict arrhythmia risk in middle-aged adults

**DOI:** 10.64898/2026.06.01.26354663

**Authors:** Qin Lu, Wen-Tao Bi, Yun-Jiu Cheng, Yu-Jie Li, Hao Tang, Li-Juan Liu

## Abstract

**Background:** Higher METS-IR has been shown to be associated with a higher risk of major adverse cardiovascular events, but data are lacking regarding cardiac arrhythmias. Objectives: The aim of this study was to assess the association between METS-IR and atrial fibrillation/flutter, ventricular arrhythmia and bradyarrhythmia.

**Methods:** Data from the Atherosclerosis Risk in Communities study spanning 1987 to 2013 was utilized for this analysis. METS-IR scores were assessed at baseline (1987-1989) and arrhythmia episodes were identified using ICD-9 codes. Multivariate-adjusted Cox proportional hazard models were constructed to evaluate the relationship between METS-IR and arrhythmia risk, with dose-response analyses conducted. In addition, we analyzed the predictive value of METS-IR for arrhythmias. Results: Over a mean follow-up of 21.9 years, 2493 cases of AF, 688 cases of bradyarrhythmia, and 1315 cases of ventricular arrhythmia were recorded. Each interquartile range increase in METS-IR was associated with a 49% higher risk of atrial fibrillation(*P*<0.001), 29% higher risk of bradyarrhythmia(*P*<0.001), and 42% higher risk of ventricular arrhythmia(*P*<0.001). After correction for relevant confounders, the METS-IR index was significantly and positively associated with the risk of new-onset atrial fibrillation, bradyarrhythmia, and ventricular arrhythmia (*P* overall<0.05, *P* for non-linearity>0.05). Most of the results of the subgroup analyses were not significantly different. The inclusion of METS-IR in the base model improves the predictive value of the risk of arrhythmogenesis. Conclusions: There is a significant association between METS-IR and increased risk of arrhythmias.

**What’s new?:**

- This study found a significant correlation between METS-IR and the risk of developing arrhythmias in a large population-based cohort. Specifically, individuals with higher METS-IR had a significantly increased risk of developing arrhythmias, even after adjusting for other cardiovascular risk factors. In addition, there was a linear relationship between METS-IR and arrhythmia.
- The ability to predict the risk of new arrhythmias was significantly improved by adding METS-IR to the basic risk prediction model. Furthermore, a visualization tool (nomogram) incorporating METS-IR was developed, which demonstrated substantial clinical net benefit in decision curve analysis, showing comparable performance to other established insulin resistance indices (TyG, SPISE).

**What Are the Clinical Implications?:**

- The identification of high-risk patients for arrhythmias using the METS-IR Index can facilitate personalized care interventions. Healthcare providers can potentially prevent arrhythmic events by addressing modifiable risk factors such as weight loss, lipid control, and improved insulin sensitivity. This proactive approach could lead to substantial public health advantages.

## Introduction

Arrhythmias have a high incidence worldwide and there are currently around 40 million patients, with as many as 7.2 million in developed countries^[1]^. Approximately seventeen million people worldwide die each year from cardiovascular disease^[2]^, and arrhythmia is a major cause of cardiovascular complications and increased risk of sudden cardiac death. Common clinical arrhythmias include atrial fibrillation/atrial flutter, bradyarrhythmia, and ventricular arrhythmia, etc. Severe arrhythmias pose a major threat to human life by increasing disability and mortality, decreasing the quality of life of patients, and incurring high medical costs. Therefore, it is crucial to identify controllable factors that can prevent or delay arrhythmias.

Insulin resistance (IR) is a pathological condition characterized by reduced tissue response to insulin stimulation, resulting in impaired glucose uptake and oxidation, decreased glycogen synthesis, and limited suppression of lipid oxidation^[3]^. Over time, IR can lead not only directly to pathological conditions such as hyperglycaemia, hypertension, hyperlipidemia, obesity, thrombotic state, elevated inflammatory markers and endothelial dysfunction, but also indirectly to metabolic diseases such as diabetes, non-alcoholic fatty liver disease, cardiovascular disease (CVD) and even death^[4]^. Currently, there are many ways to assess IR, the gold standard being the euglycaemic-hyperinsulinaemic clamp (EHC)^[5]^, which is not suitable for routine clinical assessment due to its invasive and expensive nature. To this end, some researchers have developed several indices to assess IR based on simple formulae, such as homeostasis model assessment for IR (HOMA-IR), triglyceride-glucose index (TYG index) and metabolic score for insulin resistance (METS-IR)^[6, 7]^. The METS-IR index is a novel metabolic score that assesses and quantifies insulin resistance, derived from conventional clinical indices such as fasting plasma glucose (FPG), high-density lipoprotein cholesterol (HDL-C), triglycerides (TG) and body mass index (BMI), and has been shown to have high accuracy similar to that of the EHC^[8]^. Since its development, several studies have shown that METS-IR is associated with hypertension, diabetes and coronary heart disease^[8–10]^. However, the association of METS-IR with arrhythmias is lacking in the literature.

As early as 1995 a retrospective study showed that IR was associated with sick sinus syndrome^[11]^. Some studies have shown that IR is a risk factor for arrhythmias and may lead to prolonged QT intervals^[12]^, ST-T wave alterations^[13]^, atrial fibrillation and ventricular arrhythmias, and may even lead to sudden cardiac death. Experimental and clinical findings have revealed that abnormal glucose metabolism and IR are involved in atrial electrical and structural remodeling processes and atrial fibrillation (AF) development^[14]^. Therefore, the aim of this paper is to investigate the relationship between the METS-IR index, a novel metabolic score of insulin resistance, and arrhythmias (atrial fibrillation, bradyarrhythmia, and ventricular arrhythmia) in the general middle-aged population.

## Method

### Study population

The Atherosclerosis Risk in Communities (ARIC) Study is a prospective cohort designed to investigate the causes of atherosclerosis and its clinical outcomes, and variation in cardiovascular risk factors, medical care, and disease^[15]^. The ARIC study enrolled 15,792 middle-aged (45 to 64 years old at baseline) predominantly white and African American men and women from four US communities (Forsyth County, North Carolina; Jackson, Mississippi; suburban Minneapolis, Minnesota; and Washington County, Maryland) during the period of 1987 to 1989(visit 1)^[15]^. Subsequent follow-up visits were performed in 1990 to 1992(visit 2), 1993 to 1995(visit 3), 1996 to 1998(visit 4), and 2011 to 2013(visit 5)^[16]^.

We analysed data on the ARIC Study from 1987 to 2013. For the current analysis, we finally included 14,434 participants with complete baseline data on FPG, TG, HDL-C, BMI and comprehensive follow-up data on AF, ventricular arrhythmia and bradyarrhythmia. The ARIC Study protocol was approved by the institutional review board of each participating institution and informed consent was obtained from each study participant. Cohort dataset obtained from National Institutes of Health (NIH) Coordinating Center for Biospecimen Information and Data Storage^[17]^.

### Ethical Statement

The ARIC study followed the ethical principles of the Declaration of Helsinki regarding human experimentation. All human subjects involved in the study participated voluntarily and had signed an informed consent form. The guidelines of the ethical review committee were strictly followed during the study design, implementation and data analysis. The privacy, right to information and autonomy of the subjects were ensured during the study, and the personal information of the subjects was kept strictly confidential. All data collection and analysis in this study were approved by the ethical review board.

### Ascertainment of Metabolic Score for Insulin Resistance

Venous blood samples were collected after an overnight fast (at least 8 hours) and analyzed parameters were obtained by applying different techniques. The Metabolic Insulin Resistance Score (METS-IR) has been reported as a novel and simple index for the assessment of insulin resistance, calculated as { Ln [(2 × FPG) + TG] × BMI } / [Ln (HDL-C)] ^[8]^, where FPG, TG, and HDL-C are expressed in mg/dL, and BMI is expressed in kg/m^2^.

### Data Collection at Baseline

Study participants self-reported information on age, gender, race. Educational level was categorized into less than high school, complete high school or vocational school, and college, graduate or professional school. Smoking status were classified as current, former, and never. Alcohol consumption status was classified as drinking and not drinking. Physical activity was assessed by the Modified Baecke Physical Activity Questionnaire^[18]^. Participants were asked to bring all medications to the ARIC clinic. Heart rhythm medication use was defined if participant reported taking beta-blockers (non-selective, cardio-selective, or combination), or medication for abnormal heart rhythm^[19]^. Body mass index (BMI) was calculated as weight (in kilograms) divided by the square of height (in meters). Sitting BP was measured three times using a random-zero sphygmomanometer after 5 minutes of rest. The mean of the last two measurements were used for the analysis. Total cholesterol (TC), high-density lipoprotein cholesterol (HDL-c) and triglycerides (TG) were measured using standardized enzymatic assays, and low-density lipoprotein cholesterol (LDL-C) were calculated based on Friedewald formula^[20]^. serum glucose was measured by a hexokinase/glucose-6-phosphate dehydrogenase method^[20]^. Heart rate and left ventricular hypertrophy from a typical 12-lead electrocardiogram (ECG)^[21]^. Diabetes was defined as a fasting glucose ≥126 mg/dL, non-fasting glucose ≥200 mg/dL, self-reported physician diagnosis of diabetes, or use of anti-diabetic medication^[20]^. Hypertension was defined as systolic blood pressure (BP) ≥ 140 mm Hg and/or diastolic BP ≥ 90 mm Hg, or BP medicine use in the past 2 weeks. Prevalent coronary heart disease, prevalent heart failure were defined on the basis of self-reported information.

### Ascertainment of Outcome

From the baseline visit to December 31,2013, participants were tracked for their AF, ventricular arrhythmia and bradyarrhythmia occurrence^[22]^.

Cases of new-onset arrhythmias were determined from three different sources: electrocardiograms (ECGs) conducted during study visits, examination of hospital discharge codes, and analysis of death certificates. For instance, atrial fibrillation was assessed at each study visit through a 12-lead resting ECG transmitted to the ARIC ECG Reading Center for automatic encoding using the E Marquette 12-SL program. AF was automatically detected by a computer and authenticated by a cardiologist^[23]^. Information on hospitalizations during the follow-up period was gathered through yearly follow-up phone calls and monitoring of local hospitals. Skilled abstractors collected hospital discharge codes. The reliability of detecting AF from hospital discharge codes has been demonstrated in epidemiological studies. The presence of AF was confirmed if ICD-9-CM codes 427.31 (atrial fibrillation) or 427.32 (atrial flutter) were documented. Additionally, AF was identified from death certificates if ICD-9 427.3 or ICD-10 I48 codes were listed as contributing to the cause of death^[24]^. Included in ventricular arrhythmia were ventricular tachycardia^[25]^ (ICD-9-CM code: 427.1), ventricular flutter/fibrillation (427.4, 427.41, 427.42), cardiac arrest (427.5), and sudden cardiac death. Sudden cardiac death was characterized by an underlying cardiac cause due to the exclusion of conditions clearly unrelated to cardiac arrhythmias^[19, 26]^. Bradyarrhythmia was diagnosed based on second-degree atrioventricular (AV) block (ICD-9-CM code: 426.1, 426.10), complete AV block (426.0), sick sinus syndrome (427.81), and procedures of pacemaker implantation (37.8, V45.01, V53.31)^[27]^.

### Statistical analysis

Participants were ranked in the METS-IR quartile: Quartile 1 (<35), Quartile 2 (35–41), Quartile 3 (41–48), Quartile 4 (>48). For continuous variables, the mean and standard deviation were reported if they followed a normal distribution, otherwise the median and interquartile spacing were presented. For categorical variables, frequencies and percentages were provided. The significance of differences between groups in quartile groupings of the METS-IR index was evaluated using the chi-square test (for categorical variables), the Kruskal-Wallis test (for skewed continuous variables), or ANOVA (for normally distributed continuous variables)^[4]^.

Hazard ratios (HRs) were calculated using multivariate COX risk regression models to evaluate the risk of developing atrial fibrillation, ventricular arrhythmia, and bradyarrhythmia. The analyses were adjusted for various factors such as age, sex, race, education level, alcohol use, smoking status, diabetes, heart rate, physical activity, left ventricular hypertrophy, use of cardiac rhythm medications, self-reported coronary heart disease (CHD), heart failure (HF), hypertension, and TC. Missing data for covariates were handled using multiple imputation techniques. Additionally, restricted cubic spline curves were utilized to examine potential nonlinear relationships between continuous METS-IR scores and arrhythmia risk. The linear regression model was compared with the nonlinear model using the log-likelihood ratio test. Survival analysis curves were employed to visualize the cumulative event incidence in different METS-IR groups. Furthermore, the robustness of the results was assessed through competing risk models for events and death. Subgroup analyses were conducted based on sex, age, ethnicity, smoking status, drinking status, diabetes and hypertension. Sensitivity analyses were also carried out by excluding participants with coronary heart disease and hypertension. Subject operator characteristic (ROC) curve analysis was utilized to determine if METS-IR enhanced the predictive power of the basic arrhythmia risk model. Metrics such as area under the ROC curve, c-statistics, net reclassification improvement (NRI), and integrated discrimination improvement (IDI) were calculated to quantify the improvement in each model by including METS-IR. To investigate the extent to which the association between METS-IR and arrhythmia incidence was mediated by coronary artery disease, mediation analyses were performed. A two-tailed P-value < 0.05 was defined as statistically significant.To construct visual risk prediction models and comprehensively compare the predictive performance of METS-IR with other established non-insulin-based indices (TyG, TyG-BMI, and SPISE), multivariable logistic regression analyses were performed to calculate the overall occurrence probability of arrhythmias during the follow-up. Nomograms incorporating METS-IR and other conventional risk factors were formulated to provide a quantitative tool for predicting the individual risk of incident arrhythmias. Furthermore, Decision Curve Analysis (DCA) was utilized to evaluate the clinical utility and net benefit of the predictive models across various threshold probabilities. All statistical analyses were performed in Stata software (version 17.0), SPSS (version 26.0) and R (version 4.3.2).

## Results

### Baseline characteristics of study participants

A total of 14434 individuals were enrolled in this study, with a mean age of 54.3 ± 5.8 years and a mean follow-up of 21.9 ± 4.0 years, 54.2% were female and 75.4% were white. Table 1 groups the baseline characteristics of these subjects according to quartile of the METS-IR. As METS-IR values increased, subjects had progressively higher values of FPG, TC, TG, and BMI, and progressively lower proportions of whites, alcohol drinkers, and highly educated individuals. Subjects with higher METS-IR values were more likely to have hypertension, diabetes, coronary artery disease, and heart failure, and had higher proportions of anti-hypertensive and anti-arrhythmic drug use. Importantly, individuals with higher METS-IR had a greater prevalence of arrhythmic conditions such as atrial fibrillation, ventricular arrhythmia, and bradyarrhythmia (*P* < 0.001) (Table 1 Seen in the supplementary material).

**Table 1.**
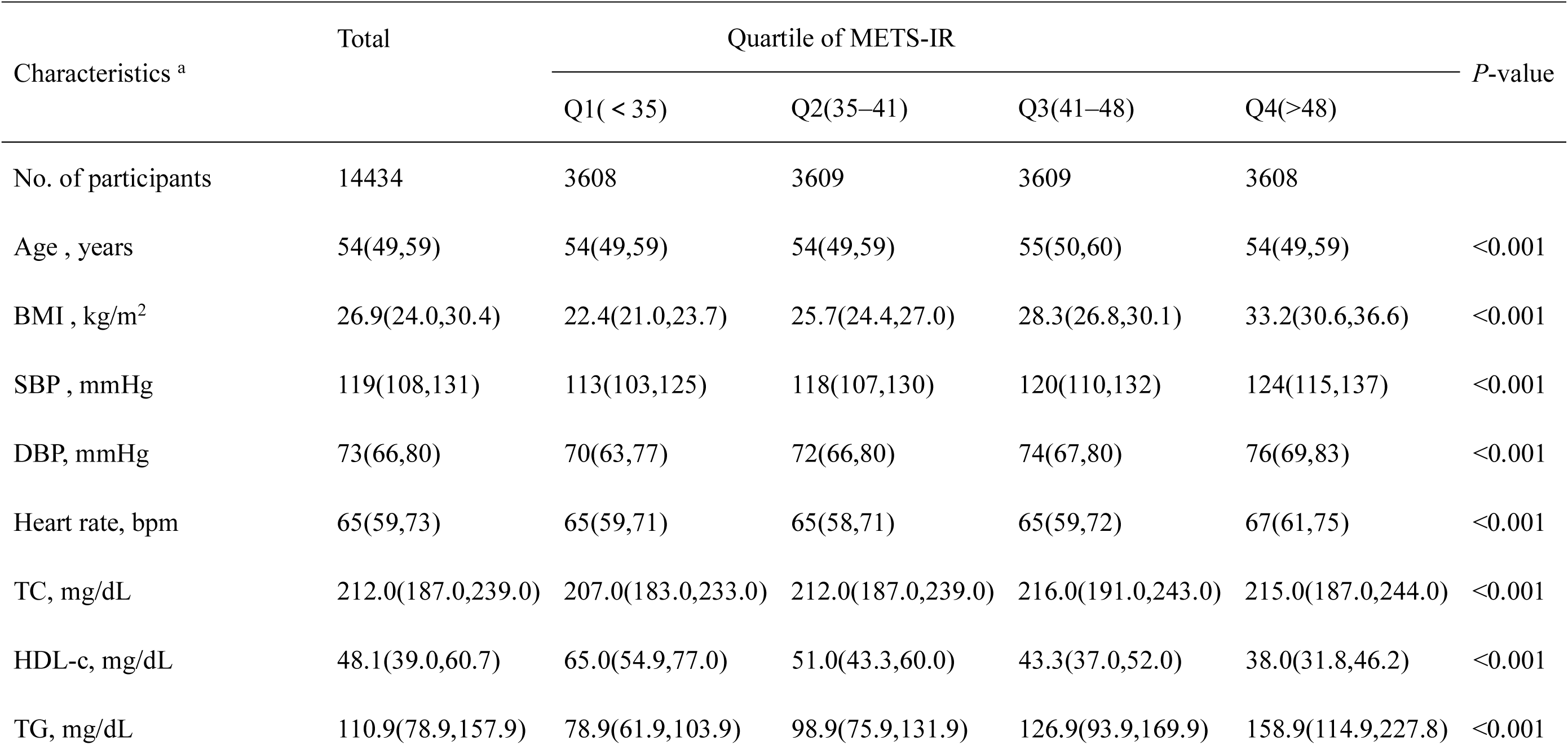
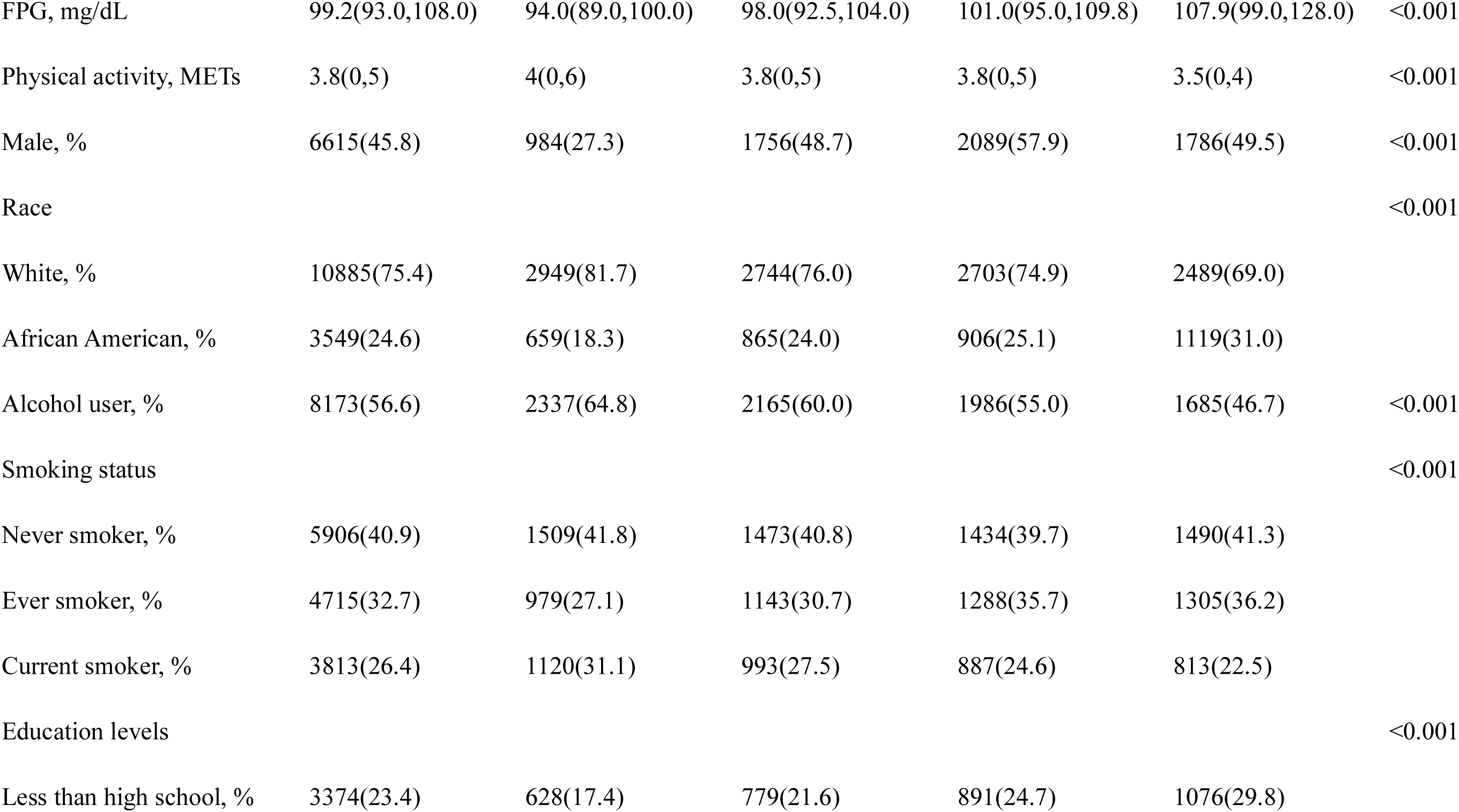
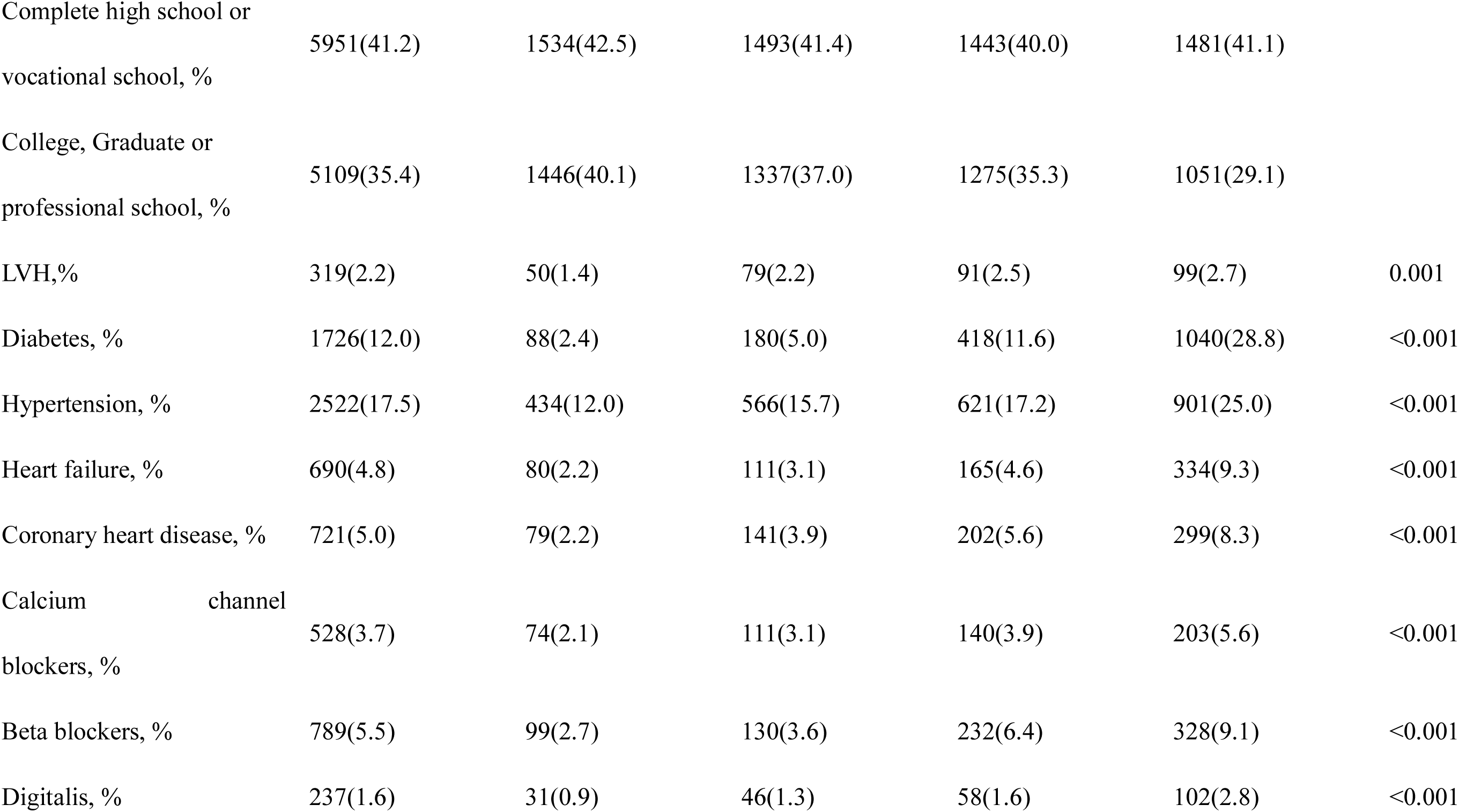
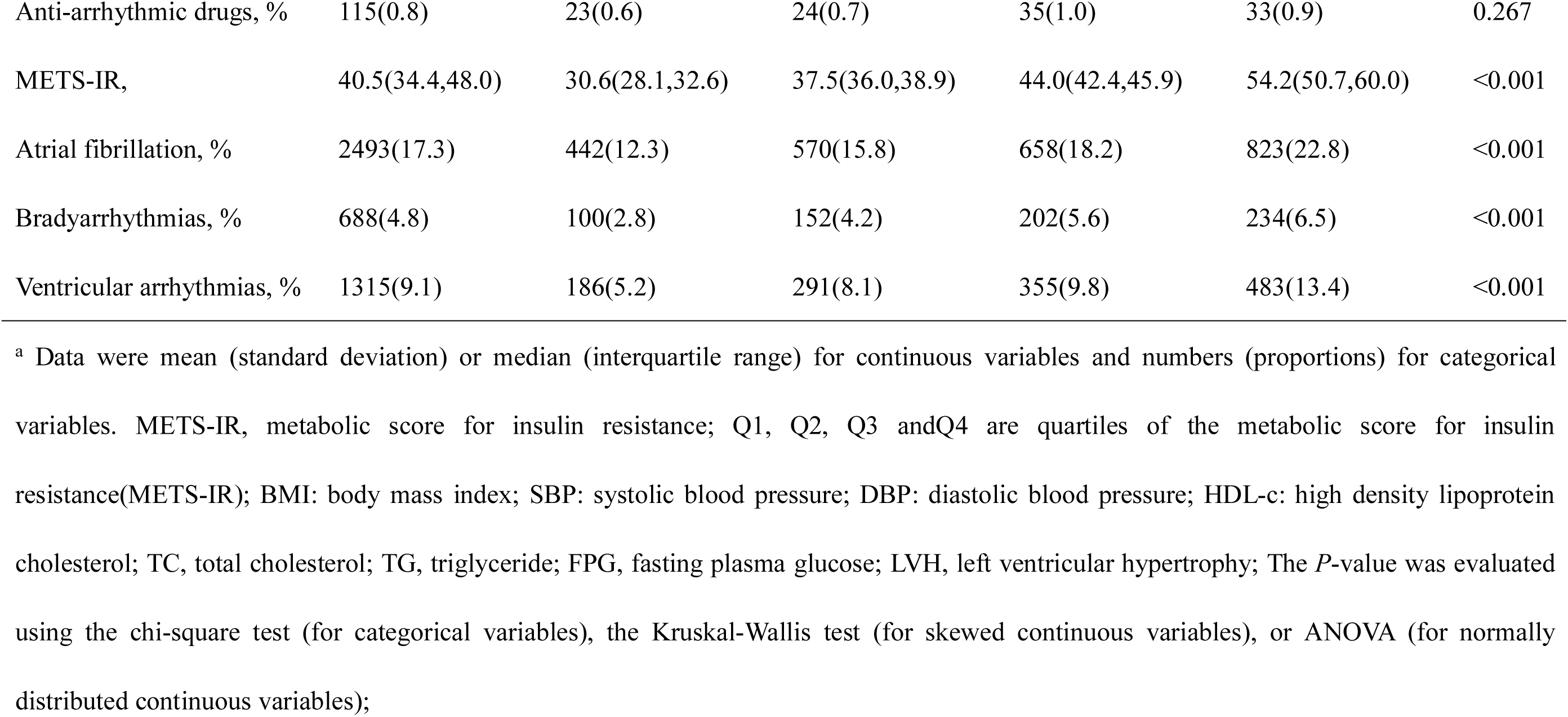
Baseline characteristics of participants by Quartile of METS-IR in ARIC 1987–1989.

### Association between baseline METS-IR and AF, ventricular arrhythmia and bradyarrhythmia

During the follow-up period, we documented 2493 incident atrial fibrillation, 688 incident bradyarrhythmia, and 1315 incident ventricular arrhythmia. The association between the novel Metabolic Score for Insulin Resistance and the risk of arrhythmias is shown in Table 2 (Table 2 Seen in the supplementary material).

**Table 2.**
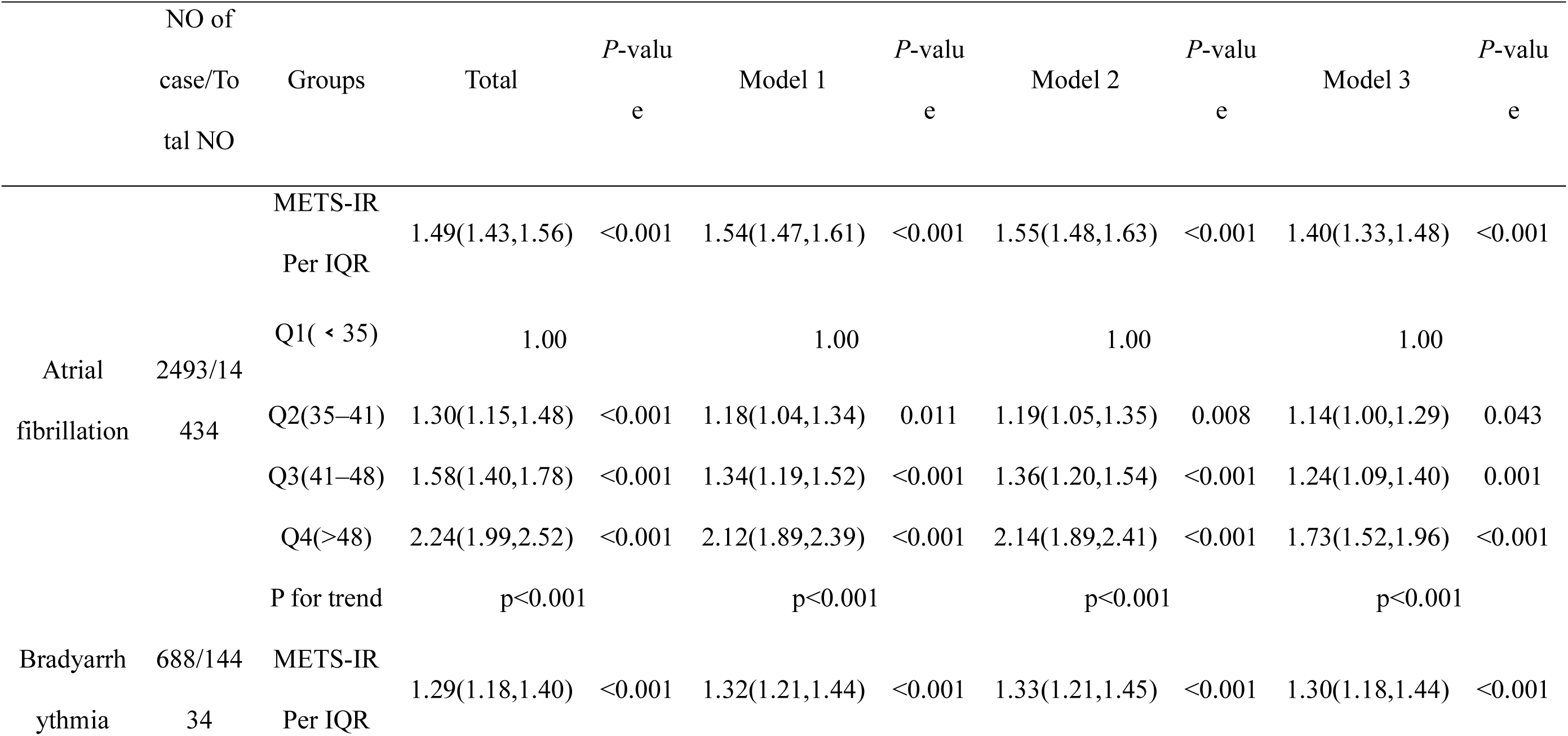
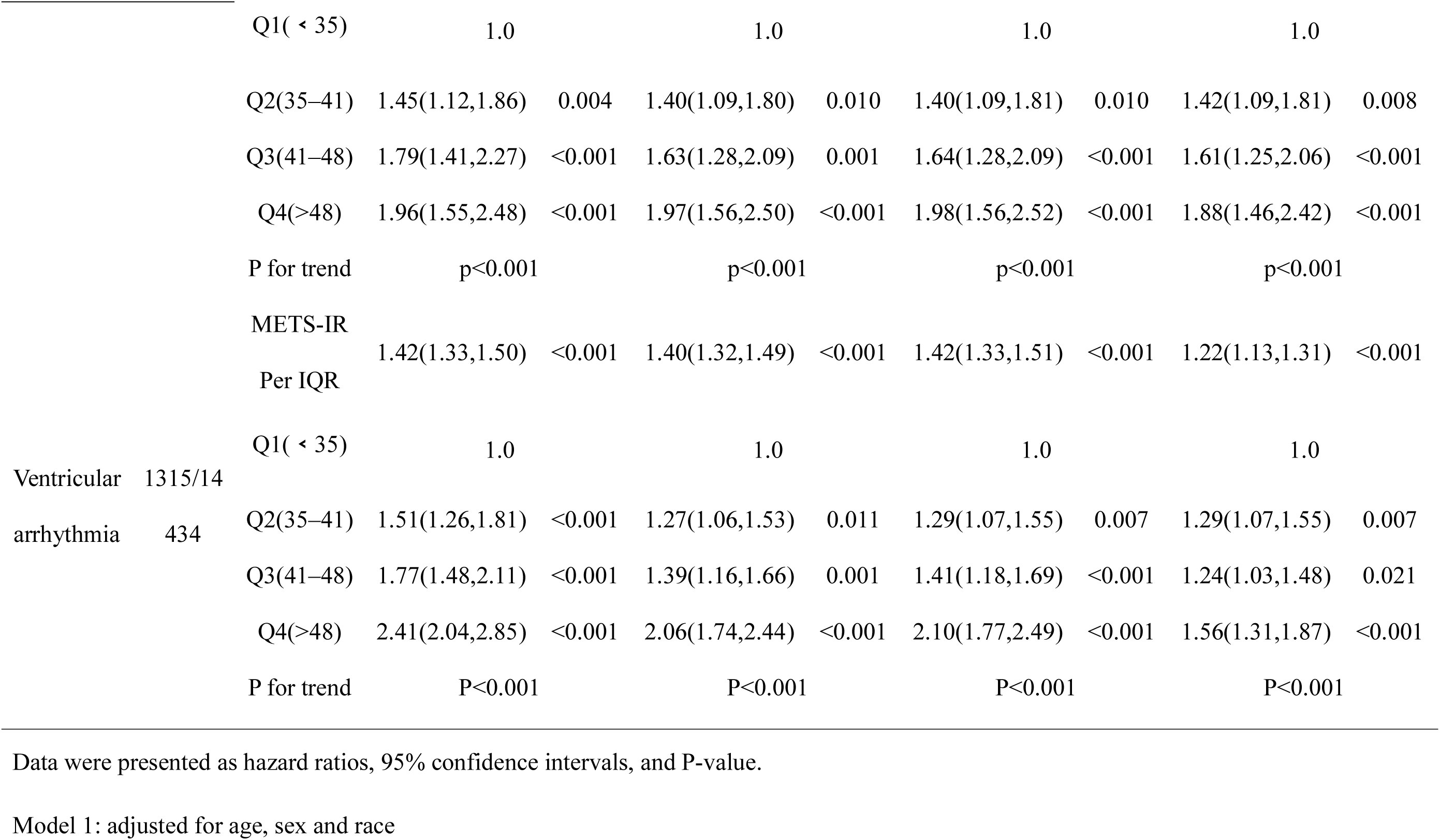
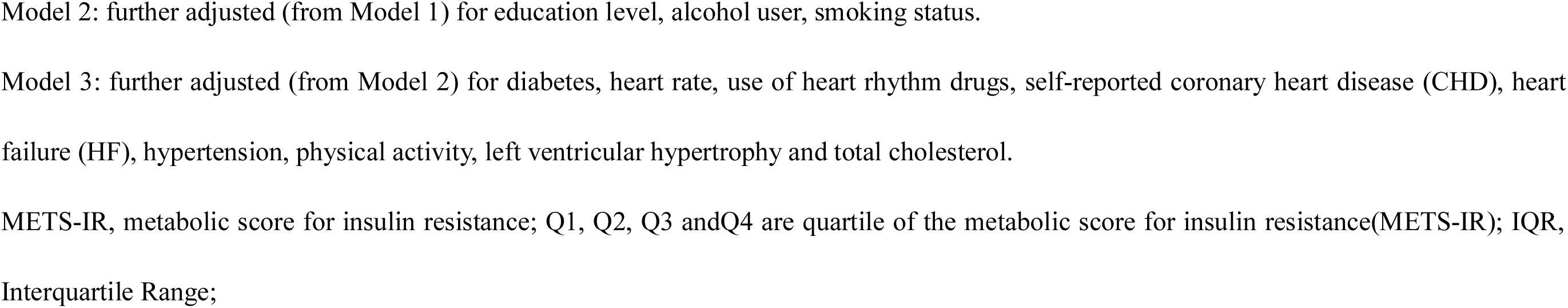
Associations between baseline METS-IR with follow-up incident atrial fibrillation/bradyarrhythmia/ventricular arrhythmia.

When analyzing METS-IR as a continuous variable, it was determined that the risk of developing AF increased by 49% for each interquartile rise (IQR) in METS-IR (HR=1.49; 95% CI, 1.43-1.56). Similar findings were noted for bradyarrhythmia and ventricular arrhythmia, showing a 29% and 41% increase in risk for each quartile increase in METS-IR (bradyarrhythmia HR=1.29, 95% CI=1.18-1.40; ventricular arrhythmia HR=1.41, 95% CI=1.33-1.50), respectively. Upon adjusting for various confounders, there were no significant differences in the risks associated with AF, bradyarrhythmia, and ventricular arrhythmia.

In quadratic subgroup analyses of the METS-IR, the risk ratios for the occurrence of ventricular arrhythmia in the Q2, Q3, and Q4 groups were 1.51 (95% CI 1.26-1.81), 1.77 (95% CI 1.48-2.11), and 2.41 (95% CI 2.04-2.85), respectively, compared with the Q1 group of the METS-IR index. After adjusting for different confounders, the risk of ventricular arrhythmia was significantly reduced in the Q2, Q3, and Q4 groups in all three models compared with the unadjusted model, with the most significant decrease in HR in the fourth (Q4) quartile group in Model 3 (HR=1.56, 95% CI=1.31-1.87), but the risk of ventricular arrhythmia, compared with the reference subgroup in Model 3 was still increased by 56%. The results for atrial fibrillation were similar to those for ventricular arrhythmia, with a 124% increased risk of developing atrial fibrillation in the fourth (Q4) quartile compared with the first (Q1) quartile of the METS-IR index (HR=2.24, 95% CI=1.99-2.52), and after adjusting for covariates, the HR for the occurrence of atrial fibrillation in Models 1, 2, and 3 was decreased compared with that when no adjustment was made for confounders, with the most significant decrease in HR values in the fourth (Q4) quartile group in model 3 (HR=1.73, 95%=1.52-1.96). For bradyarrhythmia, the fourth (Q4) quartile of the METS-IR had a 96% higher risk of developing a bradyarrhythmia than the first (Q1) quartile when not adjusted for confounders (HR=1.96, 95% CI=1.55-2.48), and after adjusting for covariates, the HR for the development of bradyarrhythmia in Models 1, 2, and 3 declined when compared with that in Model 3, which was not adjusted for confounders. compared with the decrease, which was most pronounced in model 3 (HR=1.88, 95% CI=1.46-2.42). The relationship between the METS-IR index and arrhythmic events was not substantially altered by further exclusion of participants with underlying coronary artery disease and heart failure in sensitivity analyses (Table 3 Seen in the supplementary material).

**Table 3.** Sensitivity analyses of METS-IR on atrial fibrillation/bradyarrhythmia/ventricular arrhythmia among participants after excluding participants who had CHD and HF at baseline.

|  | Atrial fibrillation | *P*-value | Bradyarrhythmia | *P*-value | Ventricular arrhythmia | *P*-value |
| --- | --- | --- | --- | --- | --- | --- |
| NO of case/Total NO | 2493/14434 |  | 688/14434 |  | 1315/14434 |  |
| METS-IR Per IQR | 1.41(1.33,1.49) | <0.001 | 1.31(1.18,1.45) | <0.001 | 1.21(1.11,1.31) | <0.001 |
| Q1( < 35) | 1.0 |  | 1.0 |  | 1.0 |  |
| Q2(35–41) | 1.12(0.98,1.28) | 0.102 | 1.41(1.08,1.84) | 0.013 | 1.18(0.96,1.44) | 0.114 |
| Q3(41–48) | 1.25(1.10,1.43) | 0.001 | 1.56(1.20,2.04) | 0.001 | 1.23(1.00,1.50) | 0.046 |
| Q4(>48) | 1.74(1.52,1.99) | <0.001 | 1.95(1.49,2.55) | <0.001 | 1.55(1.27,1.89) | <0.001 |
| P for trend | <0.001 |  | <0.001 |  | <0.001 |  |

In the restricted cubic spline diagram(Figure 1) adjusted for model 3, a linear relationship was observed between METS-IR and atrial fibrillation, bradyarrhythmia, and ventricular arrhythmia (*P* overall<0.001,*P* likelihood ratio test >0.05). METS-IR was found to be positively associated with the risk of developing these arrhythmias, with the risk increasing as METS-IR levels rose. Survival analysis curves(Figure 2) stratified by METS-IR quartile demonstrated a rising cumulative risk of AF, bradyarrhythmia, and ventricular arrhythmia with increasing METS-IR (*P* log-rank test < 0.001). Competing risk regression analyses confirmed METS-IR as a risk factor for arrhythmic events even after accounting for death as a competing event. The cumulative risk of developing these arrhythmias did not significantly change after full adjustment for covariates and continued to rise with increasing METS-IR levels (*P* for trend < 0.001) (Table 4 Seen in the supplementary material).

**Figure 1.**
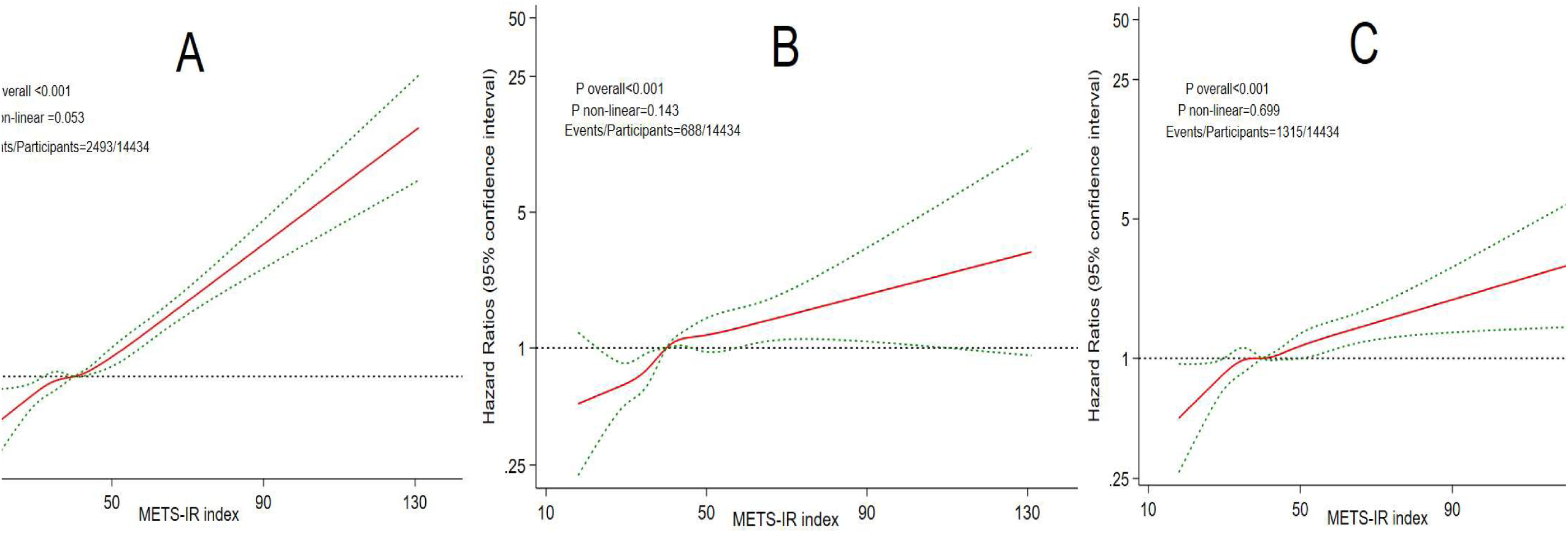
The dose-response relationships between METS-IR and the risk of atrial fibrillation (A), bradyarrhythmia (B), ventricular arrhythmia ratio (95% CI) was estimated using Cox regression mode and adjusted for age, sex, race, education level, alcohol user, smoking heart rate, use of heart rhythm drugs, self-reported hypertension, Physical activity, left ventricular hypertrophy and total cholesterol.

**Figure 2.**
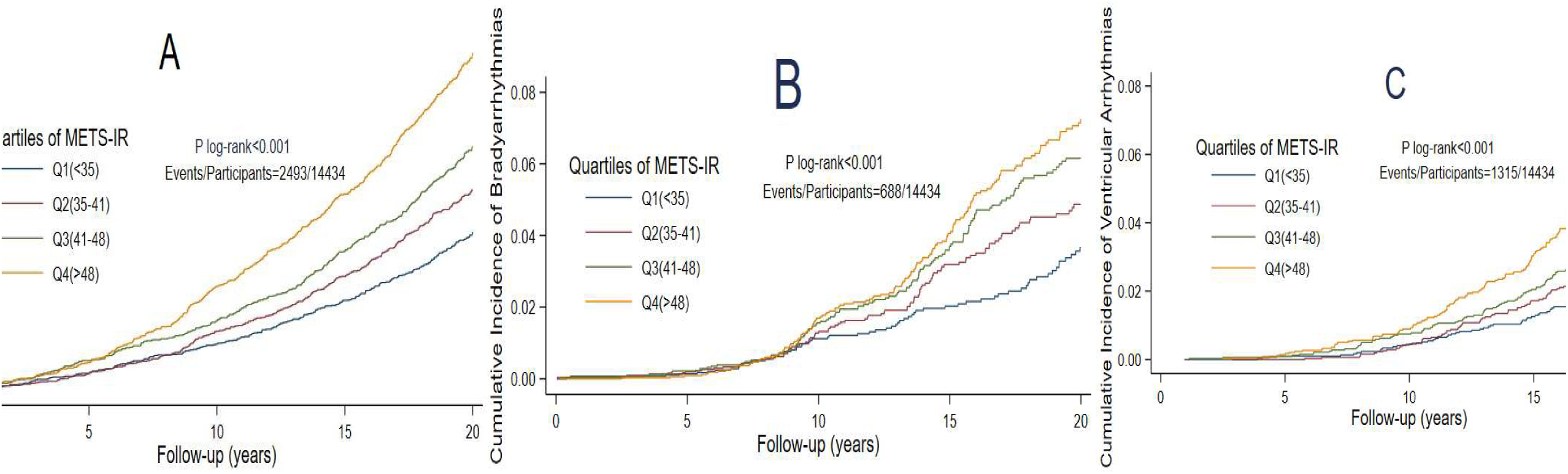
Survival analysis curve of METS-IR and the risk of incident atrial fibrillation (A), bradyarrhythmia (B), ventricular arrhythmia(ratio (95% CI) was estimated using Cox regression mode and adjusted for age, sex, race, education level, alcohol user, smoking heart rate, use of heart rhythm drugs, self-reported hypertension, Physical activity, left ventricular hypertrophy and total cholesterol.

**Table 4.** Competitive risk relationship between arrhythmic events and death.

|  | Atrial fibrillation | *P*-value | Bradyarrhythmia | *P*-value | Ventricular arrhythmia | *P*-value |
| --- | --- | --- | --- | --- | --- | --- |
| NO of case/Total NO | 2493/14434 |  | 688/14434 |  | 1315/14434 |  |
| METS-IR Per IQR | 1.37(1.30,1.45) | <0.001 | 1.27(1.16,1.40) | <0.001 | 1.19(1.11,1.28) | <0.001 |
| Q1( < 35) | 1.0 |  | 1.0 |  | 1.0 |  |
| Q2(35–41) | 1.26(1.11,1.43) | <0.001 | 1.39(1.08,1.80) | 0.010 | 1.34(1.11,1.61) | 0.002 |
| Q3(41–48) | 1.37(1.21,1.56) | <0.001 | 1.66(1.30,2.13) | <0.001 | 1.33(1.11,1.60) | 0.002 |
| Q4(>48) | 1.78(1.57,2.02) | <0.001 | 1.83(1.43,2.35) | <0.001 | 1.57(1.31,1.89) | <0.001 |
| P for trend | <0.001 |  | <0.001 |  | <0.001 |  |

Data were presented as hazard ratios, 95% confidence intervals, and P-value. Missing data of covariates were handled by the multiple imputation.
METS-IR, metabolic score for insulin resistance; Q1, Q2, Q3 and Q4 are quartiles of the metabolic score for insulin resistance(METS-IR); IQR, Interquartile Range; The Multivariate Cox regression models were adjusted for age, sex, race, education level, alcohol user, smoking status, diabetes, heart rate, use of heart rhythm drugs, self-reported hypertension, physical activity, left ventricular hypertrophy and total cholesterol.

### Layered analyses

To investigate potential variations in the relationship between METS-IR and arrhythmic events across different subgroups, participants were categorized based on socioeconomic factors and medical history (Figure 3). The findings indicated that, following adjustment for covariates, the likelihood of atrial fibrillation was 21% higher in individuals 45 to 54 years compared to those aged 55 to 64 years for atrial fibrillation events (*P*<0.001). Conversely, in the case of bradyarrhythmia events, an interaction was observed between the smoking subgroup and METS-IR, although no significant difference in event risk was noted between the subgroups. Most other subgroups did not exhibit statistically significant differences compared to the overall population, and there was no significant interaction between METS-IR and these subgroups.

**Figure 3.**
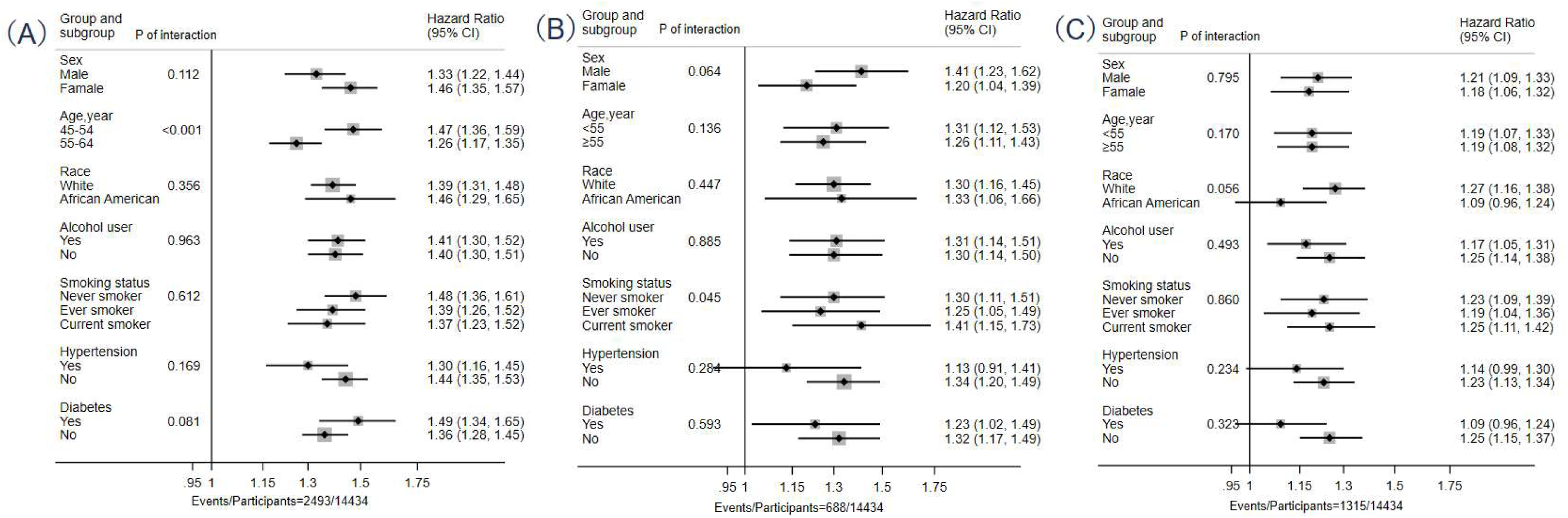
The associations between METS-IR and incident atrial fibrillation (A), bradyarrhythmia (B), ventricular arrhythmia(C) among subgroups. Hazard ratio (95% CI) was estimated using Cox regression mode and adjusted for age, sex, race, education level, alcohol user, smoking status, diabetes, heart rate, use of heart rhythm drugs, self-reported hypertension, Physical activity, left ventricular hypertrophy and total cholesterol.

### Mediation analyses

Mediation analyses showed that coronary artery disease did not play a significant role in the association of METS-IR with arrhythmia events (Table 5 Seen in the supplementary material).

**Table 5.**
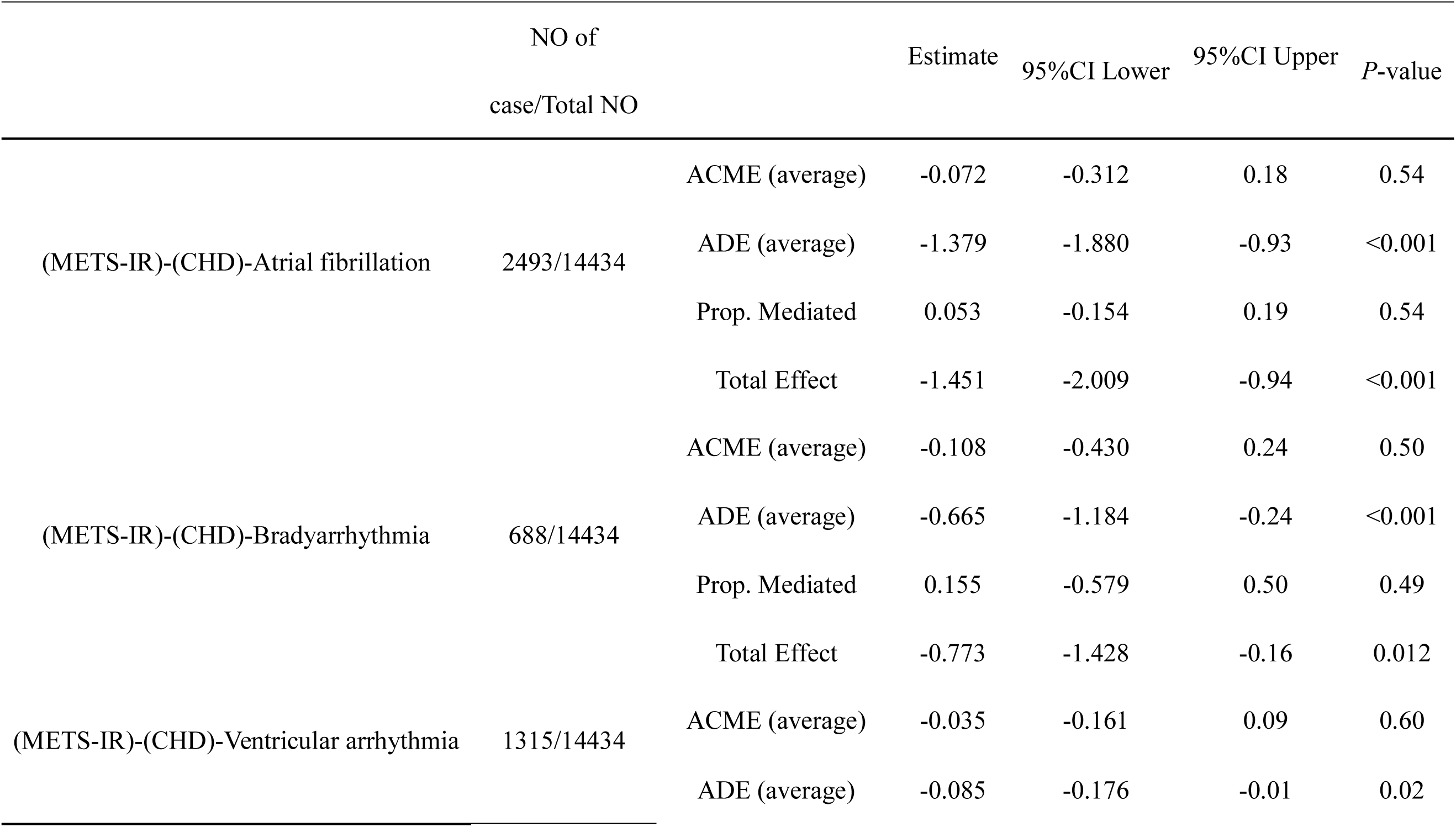
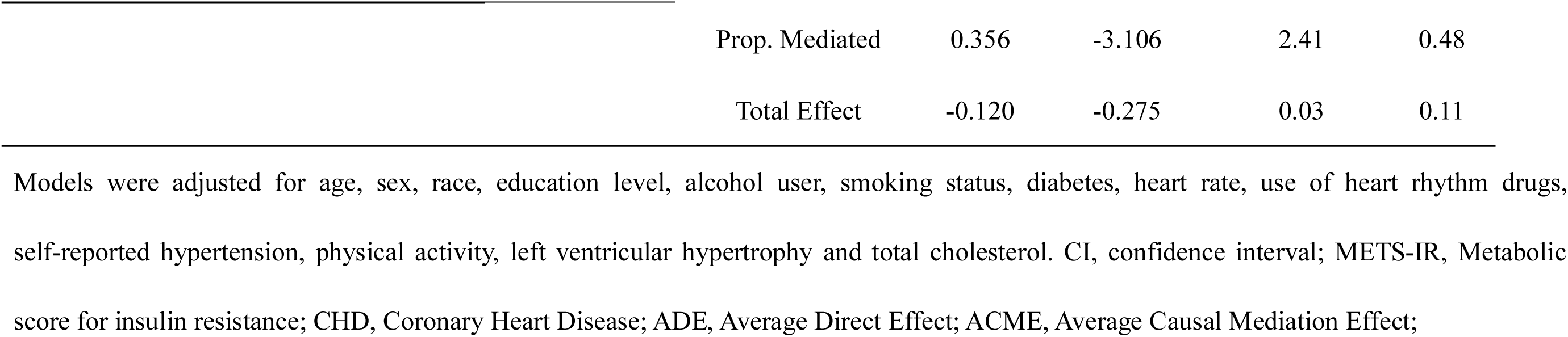
Mediation of the Association Between METS-IR and Incident atrial fibrillation / bradyarrhythmia / ventricular arrhythmia.

Predictive Performance, Clinical Utility, and Nomogram Construction Incorporating METS-IR Table 6 outlines the incremental predictive values of incorporating METS-IR into basic risk models for atrial fibrillation (AF), bradyarrhythmia, and ventricular arrhythmia. For AF, the addition of METS-IR significantly improved the discrimination ability of the model, with the C statistic increasing from 0.724 (95% CI 0.714-0.734) to 0.733 (95% CI 0.723-0.742) (P < 0.001). Similar significant improvements were observed for the continuous Net Reclassification Improvement (NRI) (3.9%; P < 0.001) and Integrated Discrimination Improvement (IDI) (0.8%; P < 0.001). Consistent trends of incremental values, though with varying magnitudes, were also observed for bradyarrhythmia and ventricular arrhythmia.

**Table 6.**
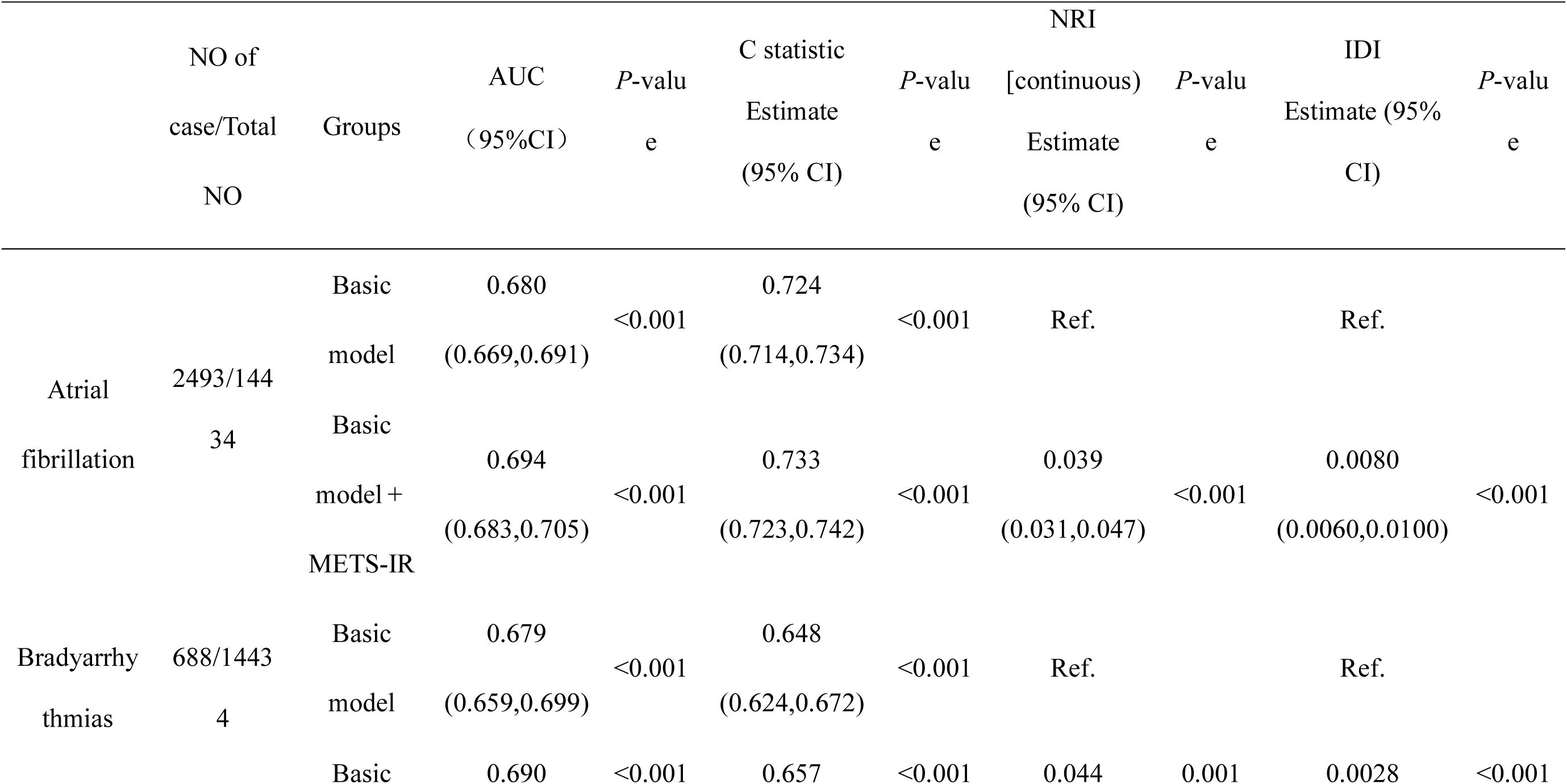
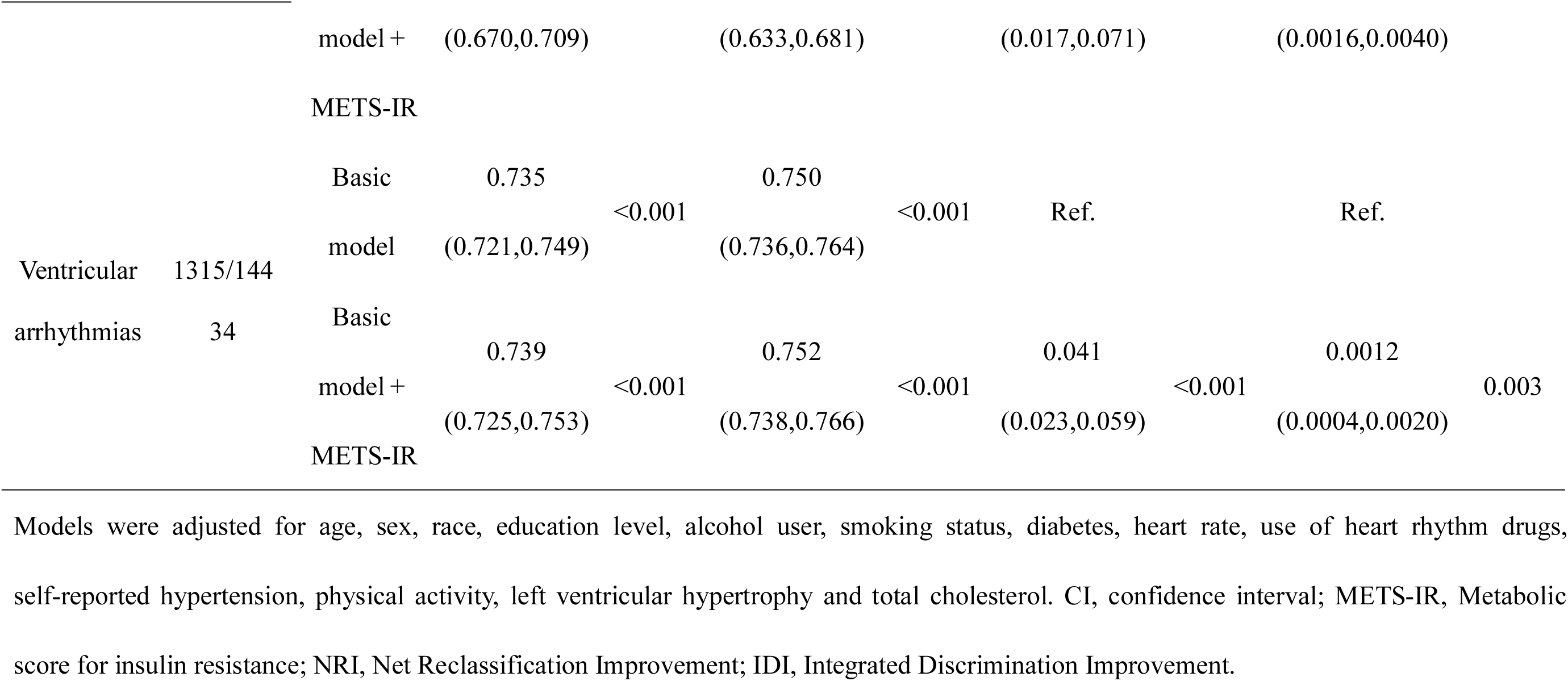
Incremental predictive value of METS-IR.

To benchmark the predictive ability of METS-IR against other well-established surrogate indices of insulin resistance, we compared its performance with the TyG index, TyG-BMI, and the SPISE index. As shown in the multi-model ROC curves (Figure 4 A-C), METS-IR demonstrated a comparable predictive performance to TyG, TyG-BMI, and SPISE across all three types of arrhythmias. Specifically, for ventricular arrhythmia, the AUC for METS-IR was 0.742, perfectly aligning with SPISE (0.742) and closely tracking TyG (0.741) and TyG-BMI (0.743).

**Figure 4.**
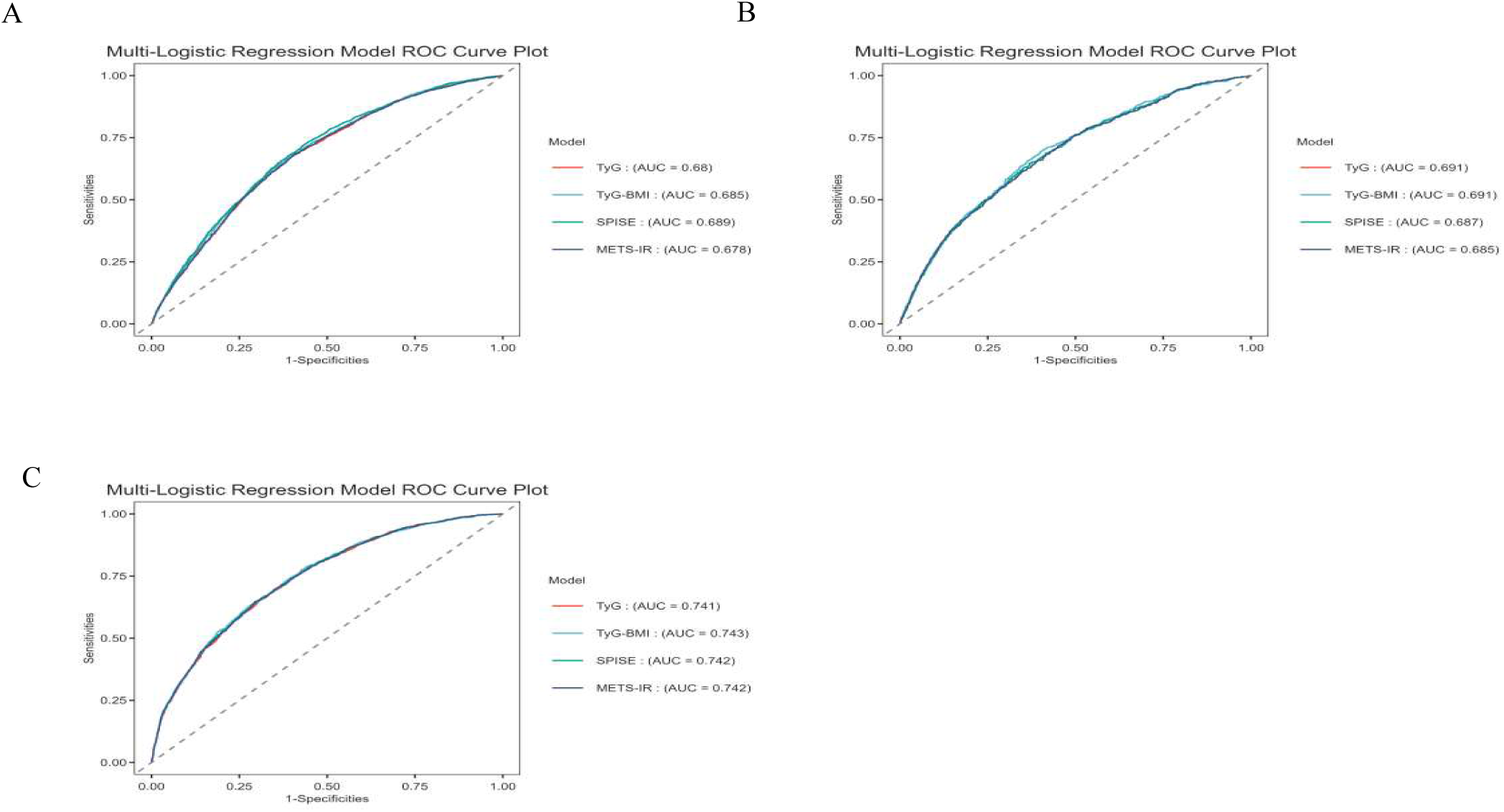
Multi-index Receiver Operating Characteristic (ROC) curves for incident arrhythmias Comparison of predictive performance among TyG, TyG-BMI, SPISE, and METS-IR for incident atrial fibrillation (A), bradyarrhythmia (B), and ventricular arrhythmia (C).

To further evaluate the practical usefulness of these indices in real-world scenarios, Decision Curve Analysis (DCA) was performed (Figure 5 A-C). The DCA curves revealed that the risk prediction model incorporating METS-IR provided a substantial positive net clinical benefit across a wide range of threshold probabilities for all three arrhythmias. Notably, the clinical net benefit trajectory of METS-IR was highly consistent and virtually overlapped with those of TyG, TyG-BMI, and SPISE, underscoring its robust non-inferiority in clinical decision-making.

**Figure 5.**
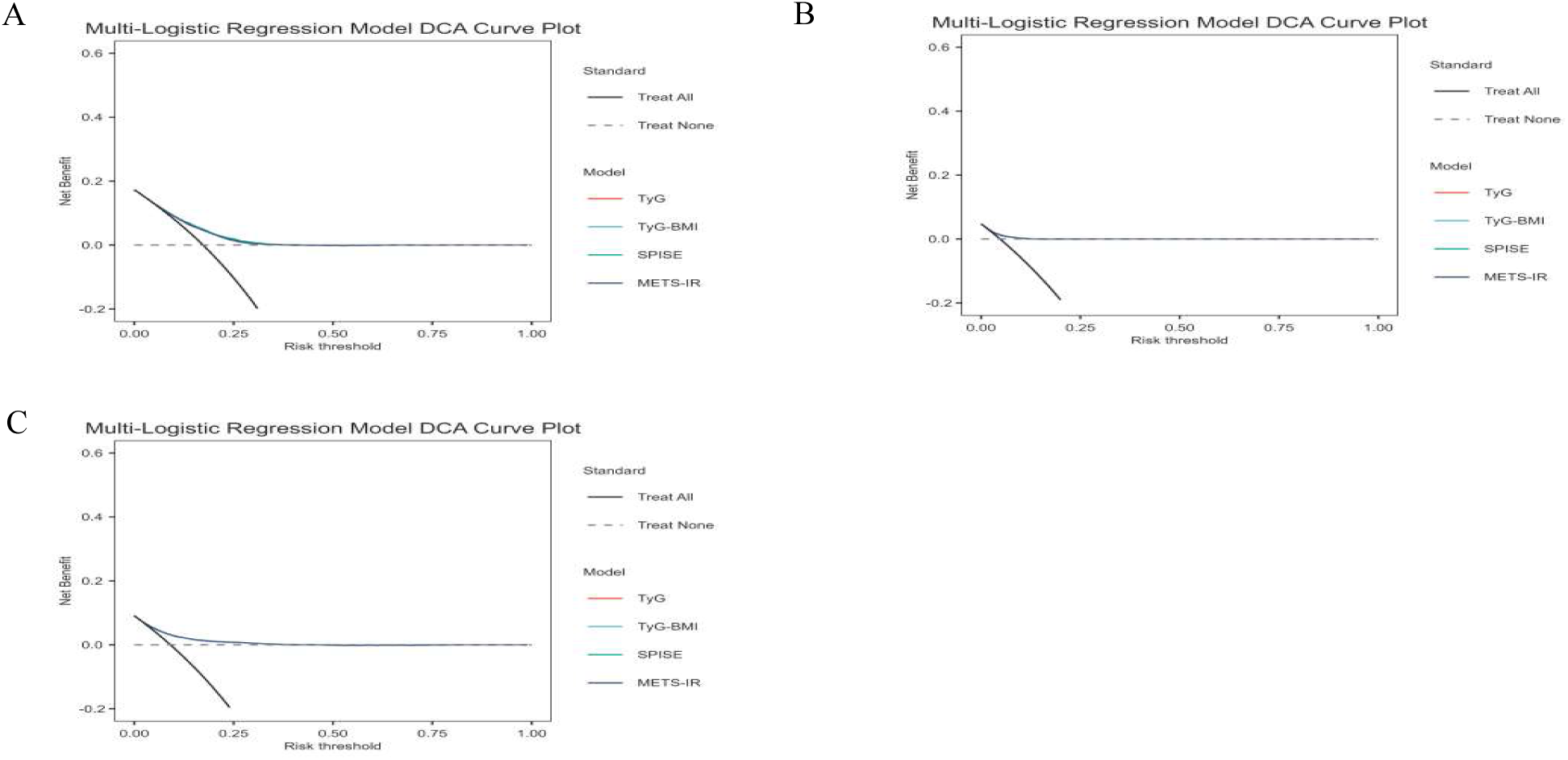
Decision Curve Analysis (DCA) for the risk prediction models The DCA curves illustrate the clinical net benefit of models incorporating TyG, TyG-BMI, SPISE, and METS-IR across different threshold probabilities for atrial fibrillation (A), bradyarrhythmia (B), and ventricular arrhythmia (C). The solid black line represents the assumption that all patients will experience the event (Treat All), and the dashed grey line represents the assumption that no patients will experience the event (Treat None).

Finally, to facilitate the translation of our findings into primary care and allow for individualized risk assessment, we developed visual risk prediction nomograms integrating METS-IR with demographic and clinical characteristics (such as age, heart rate, blood pressure, etc.) for AF, bradyarrhythmia, and ventricular arrhythmia, respectively (Figure 6 A-C). These user-friendly tools enable healthcare providers to easily calculate the individualized probability of new-onset arrhythmias, thereby guiding early and tailored preventive strategies.

**Figure 6.**
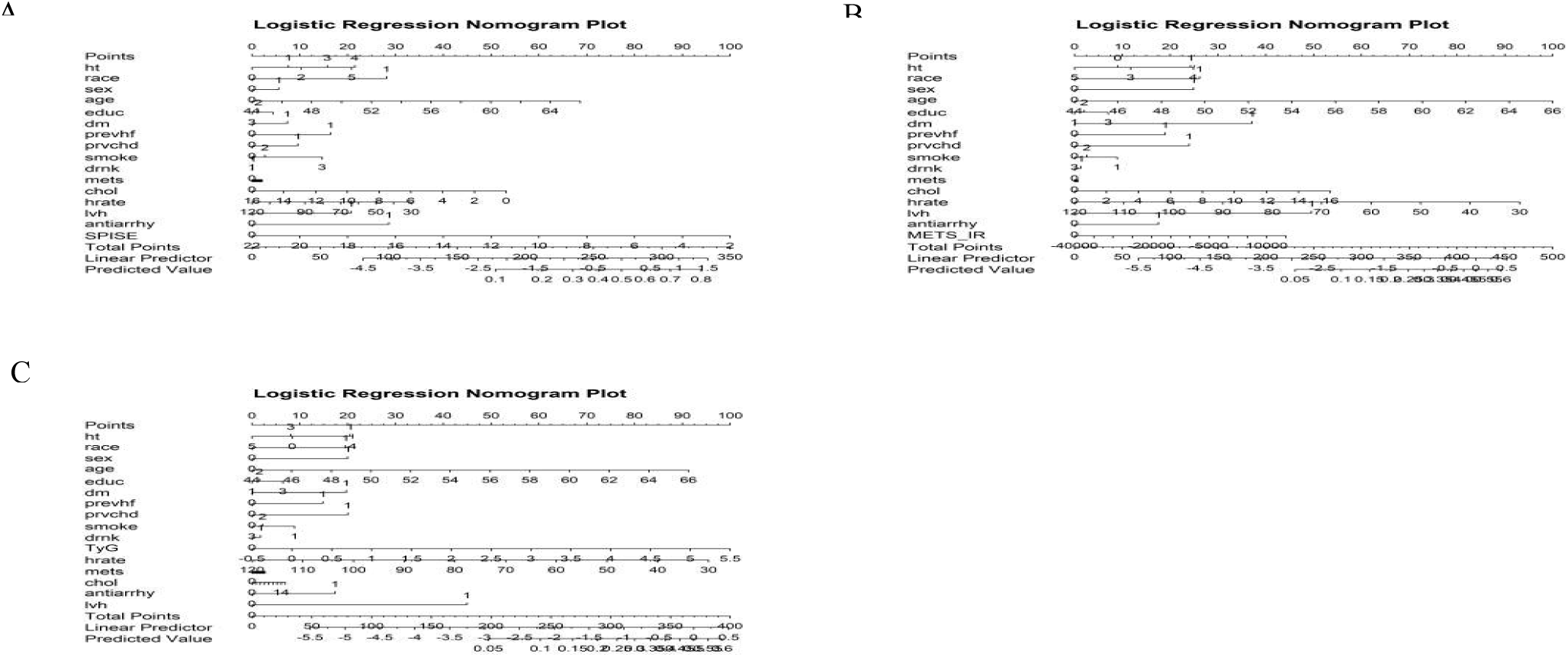
Nomograms for predicting the individual risk of incident arrhythmias Visual risk prediction nomograms incorporating METS-IR and other conventional risk factors for the incidence of atrial fibrillation (A), bradyarrhythmia (B), and ventricular arrhythmia (C). To use the nomogram, locate the patient’s value on each variable axis, draw a vertical line upwards to the "Points" axis to determine the score for that variable, sum the scores for all variables, and locate the total sum on the "Total Points" axis. Drawing a vertical line downwards to the "Predicted Value" axis gives the estimated individual probability of the arrhythmic event.

## Discussion

In this community-based prospective cohort study, we explored the relationship between METS-IR and arrhythmia, which demonstrated that METS-IR, as a continuous and categorical variable, was significantly and positively associated with the risk of arrhythmia in middle-aged Americans. And this association was not significantly related to race, gender, smoking, alcohol consumption history, or medical history. Blood glucose and cholesterol levels were much higher in METS-IR (Q4) compared to the first quartile of METS-IR (Q1). Restricted cubic spline analysis showed that METS-IR was linearly associated with arrhythmia, and the risk of arrhythmia increased with increasing METS-IR. In addition, the addition of METS-IR to the base model slightly enhanced the prediction of arrhythmia onset. To our knowledge, this is the first longitudinal cohort study to assess the relationship between baseline METS-IR and arrhythmias.

Insulin resistance (IR) is the body’s decreased response to the effects of insulin. METS-IR is an indicator of insulin resistance assessed by calculating blood glucose, lipids, and body mass index, which was validated by the glucose-high insulin clamp (EHC) test with an AUC of 0.84 (95% CI: 0.78-0.90)^[8]^. Since fasting glucose, lipids, and BMI are easily accessible, METS-IR can be used as a straightforward and precise measure of insulin resistance. Studies have revealed that a high METS-IR index is linked to hypertension, diabetes, and a higher risk of major adverse cardiovascular events^[28, 29]^. The METS-IR index is commonly used as an indicator of insulin resistance and as a predictor of various metabolic and cardiovascular issues. This study examines the relationship between the METS-IR index and cardiac arrhythmias such as atrial fibrillation (AF), ventricular arrhythmia, and bradyarrhythmia.

There are multiple indications of a relationship between insulin resistance (IR) and atrial fibrillation (AF), which have been documented in both clinical and experimental studies.Lee et al. observed that IR was linked to an elevated long-term AF risk in a non-diabetic community-based cohort^[30]^. Moreover, an investigation on non-diabetic subjects following radio-frequency ablation revealed that a higher insulin resistance index was connected to an increased likelihood of late AF recurrence^[31]^. Animal experiments have indicated that IR has the potential to raise the chances of arrhythmias by enhancing CaMKII oxidation and phospholamban phosphorylation, resulting in abnormalities in calcium balance within the cells and structural changes in the atria^[32]^. In our study, the METS-IR index was an independent risk factor for the development of AF, and we observed a trend toward a positive association between the METS-IR index and AF, with consistent results after adjusting for covariates, which is similar to the results of previous studies. In subgroup analyses, the above relationship persisted, with no significant interaction between METS-IR and subgroups other than age, suggesting that the association between METS-IR and AF risk can be applied to different combinations of subjects.

In addition, we also examined the association of the METS-IR index with ventricular arrhythmia and bradyarrhythmia, and the results were comparable. However, studies on the relationship between METS-IR and ventricular arrhythmia and bradyarrhythmia are still lacking. Some studies have shown that patients with insulin resistance experience more frequent premature ventricular beats^[33]^. In a cohort study with a 25-year follow-up, it was found that cumulative exposure to higher IR from early adulthood adversely affects left ventricular remodeling in midlife^[34]^, which may induce arrhythmias. Insulin resistance may be associated with atrioventricular block^[11]^. Our study showed that METS-IR was an independent risk factor for ventricular arrhythmias and bradyarrhythmias and that the risk of arrhythmias increased with the expansion of the index. The results remained consistent even after adjustment for variables. The association between METS-IR and arrhythmia persisted in subgroups stratified according to demographic characteristics, lifestyle, and past medical history, whereas there was no significant interaction between METS-IR and subgroups, suggesting that the association between METS-IR and the risk of ventricular arrhythmia and bradyarrhythmia generalized to most combinations of subjects in different subgroups.

The underlying mechanisms by which METS-IR is associated with arrhythmogenesis are complex and may be related to inflammation and oxidative stress. Studies have shown that insulin resistance is directly related to various inflammatory responses^[35]^, which activate the production of pro-inflammatory mediators through molecular pathways, oxidative stress, and metabolic stress. These mediators interfere with insulin signaling pathways, leading to disorders in glucose-lipid metabolism and inducing chronic inflammation, thus increasing the risk of cardiovascular disease. Furthermore, inflammation and oxidative stress have been found to contribute to cardiac electrical and structural remodeling, affecting cardiomyocytes and fibroblasts through various mechanisms. This significantly increases the risk of arrhythmic events such as tachyarrhythmias and conduction disorders. Possible mechanisms include abnormal membrane ion channel mediation^[36]^, impaired intracellular calcium handling proteins leading to intracellular calcium overload^[37]^, induction of gap junction dysfunction in cardiomyocytes^[38]^, and promotion of cardiac fibrosis^[39]^. As a result, aberrant ectopic discharges causing triggered tachyarrhythmias and the development of conduction defects may occur. In conclusion, insulin resistance increases the risk of arrhythmogenesis through inflammation and oxidative stress, as supported by the positive correlation demonstrated in our study.

In this study, the risk of atrial fibrillation was 21% higher in subjects aged 45-54 years than in those aged 55-64 years, and there was an interaction between age and METS-IR, which may be firstly related to pancreatic β-cell decline, as it has been shown that β-cells decline more rapidly in young-onset diabetes than in late-onset diabetes^[40]^, and possibly due to the narrow age range of the subjects in the study. In addition, bias may have occurred due to the narrow age range of the study population. Therefore, a broader age range and more multilevel studies could be conducted to investigate the interaction between age and the METS-IR index.

A key novelty of the current study is the direct comparison of METS-IR with other validated insulin resistance indices (TyG, TyG-BMI, and SPISE) and the subsequent construction of clinical nomograms. Previous studies have extensively validated indices like TyG in cardiovascular outcomes. In our cohort, multi-model ROC and Decision Curve Analysis (DCA) demonstrated that METS-IR possesses predictive accuracy and clinical net benefit that are highly comparable to TyG and SPISE for all three arrhythmogenic outcomes. Given that METS-IR specifically incorporates BMI alongside fasting glucose and lipid profiles, it may offer a more comprehensive reflection of the metabolic state, particularly in populations where obesity strongly drives structural atrial or ventricular remodeling. Furthermore, the METS-IR-based nomograms developed in our study provide a practical, user-friendly tool for primary physicians to visually quantify individualized arrhythmia risk using routinely available clinical parameters, bypassing the need for invasive and costly fasting insulin tests.

### Strengths and Limitations

Our research demonstrates several strengths, including being the first extensive, community-based prospective study with a long follow-up period to investigate the relationship between METS-IR and arrhythmic events. The robustness of our findings was confirmed by consistent results across different analyses and subgroups. The design of our study allowed us to explore the potential causal link between insulin resistance and arrhythmias, providing valuable insights for preventing such events. However, our research also has limitations. We did not directly measure insulin parameters in the study population, which hindered the comparison of METS-IR with direct markers of insulin resistance in predicting arrhythmia risk. Moreover, relying on admission and mortality records to identify arrhythmia occurrences may have underestimated the true incidence of such events. Additionally, our study sample only included middle-aged individuals of Caucasian and African American descent aged 45-65, which may limit the generalizability of our findings to other racial groups or age categories. Future prospective cohort studies are needed to further investigate the relationship between METS-IR and arrhythmia risk, laying the groundwork for the prevention of arrhythmic disorders.

## Conclusion

As our population continues to age, the increasing prevalence of arrhythmias poses a significant public health concern. A recent community-based study demonstrated a strong association between METS-IR and the risk of arrhythmias in middle-aged Americans, even after controlling for other variables. These results highlight the potential of METS-IR as a useful tool for early detection of arrhythmia risk, particularly in primary healthcare settings. By utilizing METS-IR as a straightforward and accessible parameter, healthcare providers may be able to intervene proactively to prevent arrhythmic events, thereby offering substantial public health benefits.

## Data Availability

The data that support the findings of this study are available from the Atherosclerosis Risk in Communities (ARIC) study. Restrictions apply to the availability of these data, which were used under license for this study. Data are available from the authors upon reasonable request and with permission of the ARIC study.

https://sites.cscc.unc.edu/aric/

## Acknowledgments

The authors thank the staff and participants of the ARIC study and Biologic Specimen and Data Repository Information Coordinating Center for their important contributions.

## Author contributions

Concept and design: L.J.L, Y.J.L, Q.L. Acquisition, analysis, or interpretation of data: Y.J.C, Q.L. Drafting of the manuscript: Q.L, E.T.B. Critical revision of the manuscript for important intellectual content: L.J.L, Y.J.C, H.T, Y.J.L. Statistical analysis: Q.L, E.T.B. All author contributed to the article and approved the submitted version.

## Data availability

Data are available upon reasonable request.

## References

[1] 李丽, 王铭铭, 孙环. 不同人群心律失常情况调查及影响因素. 中国卫生工程学. 2021. 20(01): 53–55.

[2] Moran AE, Roth GA, Narula J, Mensah GA. 1990-2010 global cardiovascular disease atlas. Glob Heart. 2014. 9(1): 3–16.

[3] Ormazabal V, Nair S, Elfeky O, Aguayo C, Salomon C, Zuiga FA. Association between insulin resistance and the development of cardiovascular disease. Cardiovasc Diabetol. 2018. 17(1): 122.

[4] Wang Z, Xie J, Wang J, Feng W, Liu N, Liu Y. Association Between a Novel Metabolic Score for Insulin Resistance and Mortality in People With Diabetes. Front Cardiovasc Med. 2022. 9: 895609.

[5] DeFronzo RA, Tobin JD, Andres R. Glucose clamp technique: a method for quantifying insulin secretion and resistance. Am J Physiol. 1979. 237(3): E214–23.

[6] Matthews DR, Hosker JP, Rudenski AS, Naylor BA, Treacher DF, Turner RC. Homeostasis model assessment: insulin resistance and beta-cell function from fasting plasma glucose and insulin concentrations in man. Diabetologia. 1985. 28(7): 412–9.

[7] Ding X, Wang X, Wu J, Zhang M, Cui M. Triglyceride-glucose index and the incidence of atherosclerotic cardiovascular diseases: a meta-analysis of cohort studies. Cardiovasc Diabetol. 2021. 20(1): 76.

[8] Bello-Chavolla OY, Almeda-Valdes P, Gomez-Velasco D, et al. METS-IR, a novel score to evaluate insulin sensitivity, is predictive of visceral adiposity and incident type 2 diabetes. Eur J Endocrinol. 2018. 178(5): 533–544.

[9] Liu XZ, Fan J, Pan SJ. METS-IR, a novel simple insulin resistance indexes, is associated with hypertension in normal-weight Chinese adults. J Clin Hypertens (Greenwich). 2019. 21(8): 1075–1081.

[10] Zhang M, Liu D, Qin P, et al. Association of metabolic score for insulin resistance and its 6-year change with incident type 2 diabetes mellitus. J Diabetes. 2021. 13(9): 725–734.

[11] Wasada T, Katsumori K, Hasumi S, et al. Association of sick sinus syndrome with hyperinsulinemia and insulin resistance in patients with non-insulin-dependent diabetes mellitus: report of four cases. Intern Med. 1995. 34(12): 1174–7.

[12] Kahn JK, Sisson JC, Vinik AI. QT interval prolongation and sudden cardiac death in diabetic autonomic neuropathy. J Clin Endocrinol Metab. 1987. 64(4): 751–4.

[13] Molon G, Costa A, Bertolini L, et al. Relationship between abnormal microvolt T-wave alternans and poor glycemic control in type 2 diabetic patients. Pacing Clin Electrophysiol. 2007. 30(10): 1267–72.

[14] Tang Q, Guo XG, Sun Q, Ma J. The pre-ablation triglyceride-glucose index predicts late recurrence of atrial fibrillation after radiofrequency ablation in non-diabetic adults. BMC Cardiovasc Disord. 2022. 22(1): 219.

[15] The Atherosclerosis Risk in Communities (ARIC) Study: design and objectives. The ARIC investigators. Am J Epidemiol. 1989. 129(4): 687–702.

[16] Lu Y, Ballew SH, Tanaka H, et al. 2017 ACC/AHA blood pressure classification and incident peripheral artery disease: The Atherosclerosis Risk in Communities (ARIC) Study. Eur J Prev Cardiol. 2020. 27(1): 51–59.

[17] Giffen CA, Carroll LE, Adams JT, Brennan SP, Coady SA, Wagner EL. Providing Contemporary Access to Historical Biospecimen Collections: Development of the NHLBI Biologic Specimen and Data Repository Information Coordinating Center (BioLINCC). Biopreserv Biobank. 2015. 13(4): 271–9.

[18] Parvathaneni K, Surapaneni A, Ballew SH, et al. Association Between Midlife Physical Activity and Incident Kidney Disease: The Atherosclerosis Risk in Communities (ARIC) Study. Am J Kidney Dis. 2021. 77(1): 74–81.

[19] Zhao D, Post WS, Blasco-Colmenares E, et al. Racial Differences in Sudden Cardiac Death. Circulation. 2019. 139(14): 1688–1697.

[20] Liao LZ, Zhang SZ, Li WD, et al. Serum albumin and atrial fibrillation: insights from epidemiological and mendelian randomization studies. Eur J Epidemiol. 2020. 35(2): 113–122.

[21] Desai CS, Ning H, Lloyd-Jones DM. Competing cardiovascular outcomes associated with electrocardiographic left ventricular hypertrophy: the Atherosclerosis Risk in Communities Study. Heart. 2012. 98(4): 330–4.

[22] Elliott AD, Linz D, Mishima R, et al. Association between physical activity and risk of incident arrhythmias in 402406 individuals: evidence from the UK Biobank cohort. Eur Heart J. 2020. 41(15): 1479–1486.

[23] Alonso A, Agarwal SK, Soliman EZ, et al. Incidence of atrial fibrillation in whites and African-Americans: the Atherosclerosis Risk in Communities (ARIC) study. Am Heart J. 2009. 158(1): 111–7.

[24] Decker JJ, Norby FL, Rooney MR, et al. Metabolic Syndrome and Risk of Ischemic Stroke in Atrial Fibrillation: ARIC Study. Stroke. 2019. 50(11): 3045–3050.

[25] Norby FL, Alonso A, Rooney MR, et al. Association of Ventricular Arrhythmias With Dementia: The Atherosclerosis Risk in Communities (ARIC) Study. Neurology. 2021. 96(6): e926–e936.

[26] Zhang Y, Guallar E, Ashar FN, et al. Association between mitochondrial DNA copy number and sudden cardiac death: findings from the Atherosclerosis Risk in Communities study (ARIC). Eur Heart J. 2017. 38(46): 3443–3448.

[27] Alonso A, Jensen PN, Lopez FL, et al. Association of sick sinus syndrome with incident cardiovascular disease and mortality: the Atherosclerosis Risk in Communities study and Cardiovascular Health Study. PLoS One. 2014. 9(10): e109662.

[28] Cheng H, Yu X, Li YT, et al. Association between METS-IR and Prediabetes or Type 2 Diabetes Mellitus among Elderly Subjects in China: A Large-Scale Population-Based Study. Int J Environ Res Public Health. 2023. 20(2).

[29] Qian T, Sheng X, Shen P, Fang Y, Deng Y, Zou G. Mets-IR as a predictor of cardiovascular events in the middle-aged and elderly population and mediator role of blood lipids. Front Endocrinol (Lausanne). 2023. 14: 1224967.

[30] Lee Y, Cha SJ, Park JH, et al. Association between insulin resistance and risk of atrial fibrillation in non-diabetics. Eur J Prev Cardiol. 2020. 27(18): 1934–1941.

[31] Wang Z, Wang YJ, Liu ZY, et al. Effect of Insulin Resistance on Recurrence after Radiofrequency Catheter Ablation in Patients with Atrial Fibrillation. Cardiovasc Drugs Ther. 2023. 37(4): 705–713.

[32] Maria Z, Campolo AR, Scherlag BJ, Ritchey JW, Lacombe VA. Dysregulation of insulin-sensitive glucose transporters during insulin resistance-induced atrial fibrillation. Biochim Biophys Acta Mol Basis Dis. 2018. 1864(4 Pt A): 987–996.

[33] Provotorov VM, Glukhovski ML. [Ventricular extrasystole in patients with metabolic syndrome]. Klin Med (Mosk). 2010. 88(1): 29–31.

[34] Kishi S, Gidding SS, Reis JP, et al. Association of Insulin Resistance and Glycemic Metabolic Abnormalities With LVStructure and Function inMiddle Age: The CARDIA Study. JACC Cardiovasc Imaging. 2017. 10(2): 105–114.

[35] Rehman K, Akash MS. Mechanisms of inflammatory responses and development of insulin resistance: how are they interlinked. J Biomed Sci. 2016. 23(1): 87.

[36] Drew BJ, Ackerman MJ, Funk M, et al. Prevention of torsade de pointes in hospital settings: a scientific statement from the American Heart Association and the American College of Cardiology Foundation. Circulation. 2010. 121(8): 1047–60.

[37] Wit AL. Afterdepolarizations and triggered activity as a mechanism for clinical arrhythmias. Pacing Clin Electrophysiol. 2018 .

[38] De Jesus NM, Wang L, Herren AW, et al. Atherosclerosis exacerbates arrhythmia following myocardial infarction: Role of myocardial inflammation. Heart Rhythm. 2015. 12(1): 169–78.

[39] Frangogiannis NG. Cardiac fibrosis. Cardiovasc Res. 2021. 117(6): 1450–1488.

[40] Magliano DJ, Sacre JW, Harding JL, Gregg EW, Zimmet PZ, Shaw JE. Young-onset type 2 diabetes mellitus - implications for morbidity and mortality. Nat Rev Endocrinol. 2020. 16(6): 321–331.

